# The use of a deep learning model in the histopathological diagnosis of actinic keratosis: A case control accuracy study

**DOI:** 10.1101/2023.11.20.23298649

**Authors:** J. Balkenhol, M. Schmidt, T. Schnauder, J. Langhorst, J. Le’Clerc Arrastia, D. Otero Baguer, G. Gilbert, L. Schmitz, T. Dirschka

## Abstract

Actinic Keratosis (AK) is a frequent dermatological diagnosis which contributes to a large proportion of routine dermatopathology. A current development in histopathology is in the digitization of specimens by creating whole slide images (WSI) with slide scanners. Deep Learning Models (DLM) have been introduced to radiology or pathology for image recognition but dermatopathology lacks available solutions. Building on previous work about skin pathologies, this paper proposes a DLM following the U-Net architecture to detect AK in histopathological samples. In total, 297 histopathological slides (269 with AK and 28 without AK) have been retrospectively selected. They were randomly assigned to training, validation, and testing groups. Performance was evaluated by conducting a Case Control Accuracy Study on three levels of granularity. The DLM model achieved an overall accuracy of 99.13% on the WSI level, 99.02% on the patch level and an intersection over union (IoU) of 83.88%. The proposed DLM reliably recognizes AK in histopathological images, supporting the implementation of DLMs in dermatopathology practice. Given existing technical capabilities and advancements, DLMs could have a significant influence on dermatopathology routine in the future.

## 1 Introduction

Actinic keratoses (AK) represent early in-situ squamous cell carcinomas of the skin which may have the potential to progress into invasive skin cancer [1]. AK has an ever-growing incidence and is one of the most common dermatological presentations [2], [3]. AK is predominantly caused by chronic ultraviolet ray exposure causing keratinocyte damage [2], [4]. Patients with AK often have a high treatment burden due to multiple lesions and a high incidence of relapse [5], [6]. Histologically, AK is characterized by atypical pleomorphic keratinocytes, limited to the epidermis with intact basal membrane [7]. Proliferative keratinocytes extend from basal cells of the epidermis and show variable extents of upward and downward-directed growth [8], [9]. Although AKs are not usually biopsied for diagnosis or treatment [24], they are very common in clinical and histological practice. In a retrospective analysis of the years 2018-2022 from the laboratory of which the specimens for this work were obtained, AK represented 19.64% of the total cases. Dermatopathologists are often limited by diagnostic uncertainty. Practitioners use symptoms, examination signs and results of investigations, using Fricke’s ‘Data-Information-Knowledge-Wisdom pyramid’ to aid diagnostic decision making [10]. This is inevitably limited by human capacity due to processing speed, performance variability and data storage ability. Artificial neural networks (ANN), which resemble the architecture of biological neural networks, have been developed as computational solutions to address the limitations of human capacity [11]. ANNs consist of multiple layers of which each contains a variable number of neurons. Three layers exist: the input layer where all information is inserted; variable numbers of hidden layers which filter the inserted data for patterns; and, finally, the output layer. Deep neural networks (DNN) are a subcategory of ANN with strengths in image data classification and segmentation. Advanced DNN architecture, ‘U-Net’ was specifically designed for processing biomedical image data. This has significantly reduced the amount of annotated training data that is required [12]. The usability of histological image data has also improved in terms of availability and quality. Slide scanners generate interactive digital pictures with high resolution from glass histology slides. The process is increasingly common in dermatopathology and enables use of DNN applications in clinical practice [13]. The U-Net architecture has been used successfully for histological analysis of breast and lung cancer [14]–[16]. While deep learning models (DLMs) for dermatopathology have been proposed [17]–[21], there is limited research in this field. Olsen et al. have shown that artificial intelligence (AI) can accurately classify ∼99% of seborrheic keratosis (SK), dermal nevus and nodal basal cell carcinoma [19]. Previous studies corroborate these findings by showing 96.4% accuracy in recognizing BCC and comparable performance for Morbus Bowen (MB) and SK [21], [22]. Moreover, approaches for assisting dermatopathologists have been demonstrated with evaluation of margin sections [27]. However, significant challenges remain in transferring AI from research into clinical practice [23]. For example, there are high hardware costs and an absence of commercially available AI software. In the first instance, DLMs must gain an evidence base which proves them to be effective in diagnosing a wide range of dermatological conditions. Classification of AK was chosen for this study as it is a frequent diagnosis in dermatopathology routine. The main objective of this case control accuracy study was to evaluate a DLMs ability to recognize exact areas of AK in histological samples.

## 2 Materials and Methods

The study was approved by the University Witten-Herdecke’s ethics committee (Proposal-No. 105/2022). In total, 297 histopathological slides (269 with AK and 28 without) were retrospectively selected from the CentroDerm Clinic GmbH (Wuppertal, Germany) database, in the period 2018 to 2022. The biopsies were performed in clinical routine. All H&E-stained slides were initially analyzed by one dermatopathologist who diagnosed AK. Two dermatopathologists applied the inclusion and exclusion criteria and selected appropriate samples for the training group from AKs of any variant, classification, excision technique, location age and sex. Sex of participants was assigned based on self-report. None of the patients have been particularly examined for sex in terms of body characteristics or chromosomal genotype.

Histological inclusion criteria:

a. Complete preservation and visibility of the epidermis
b. Presence of the stratum papillare with at least double the thickness of the epidermal layer overlying it
c. Visibility of stratum papillare over at least 50% of the epidermis length

Exclusion criteria:

a. Co-pathology

Also included were 28 specimens with healthy tissue from tumor margin sections or Burrow triangle skin from flap surgery. They were used to train the DLM to recognize healthy epidermis. Cases were randomly assigned to training and test groups and model performance on healthy slides not utilized for training was tested. All slides were digitized with the Hamamatsu Nanozoomer S210 slide scanner. The scanner creates an interactive photograph referred to as a whole slide image (WSI) in the file format .ndpi. The WSI can be digitally viewed with software and navigation is possible in three dimensions. Third dimension navigation corresponds to magnification in the microscope. For the training of the DLM, the areas in the WSI with AK were marked, referred to as annotation. The areas with AK were manually annotated in the online platform DigiPathViewer which is being developed by parts of this research group. The annotations included any AK plus hair follicle extension. Annotations were confirmed by at least one board-certified dermatopathologist.

### 2.1 DLM architecture

The DLM was developed by using a multi-step procedure to detect and segment AK in WSI of histopathological slides (Figure 1). WSI have large resolution, resulting in large memory requirements during processing. Areas with tissue were isolated to avoid unnecessary evaluation of the background area. A combination of classic image filters, morphological operations, and a threshold method were used for this task. Secondly, the tissue areas were divided into smaller image patches of size 1024px by 1024px at a magnification level of 10x by a greedy partitioning algorithm. The patches had an overlap of at least 512px in all directions. At this point, the image patches were processed by the DLM. We used a DNN based on a modified version of the established U-Net architecture [12] for the segmentation of AK. Variants of this network have already been analyzed on detection of BCC [21], SK, Bowen’s disease [22] and melanoma metastases in lymph nodes [18]. The DLM outputs a value in the range (0-1) for each pixel of an analyzed patch. This value can be interpreted as a probability score, that a pixel belongs to a region with AK. The probability outputs were then combined by a mathematical model to calculate the final AK probability for each pixel of the entire WSI. The probability values can be displayed as a heatmap directly on the WSI (Figure 2). Furthermore, all pixels that had a probability value above a set threshold were assigned the class AK. Values below this threshold are considered normal. The result is a segmentation map of the AK areas on the WSI. This marks the output of the processing pipeline. In this study, the segmentation map created by the DLM was compared with the annotations provided by the dermatopathologists.

**Figure 1.**
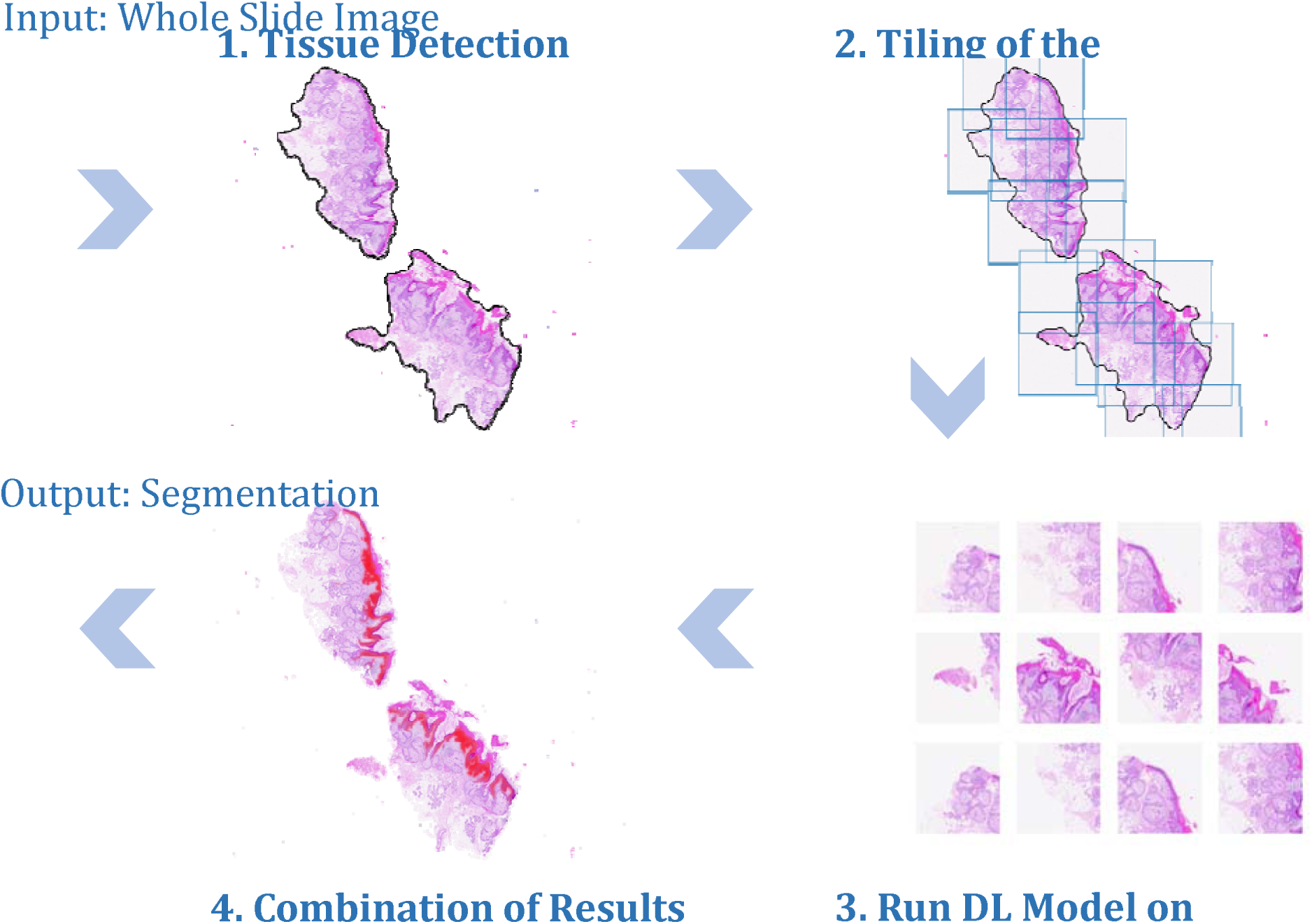
“Multi-step segmentation”. Processing steps used to generate segmentation masks that highlight the AK regions in a WSI.

**Figure 2.**
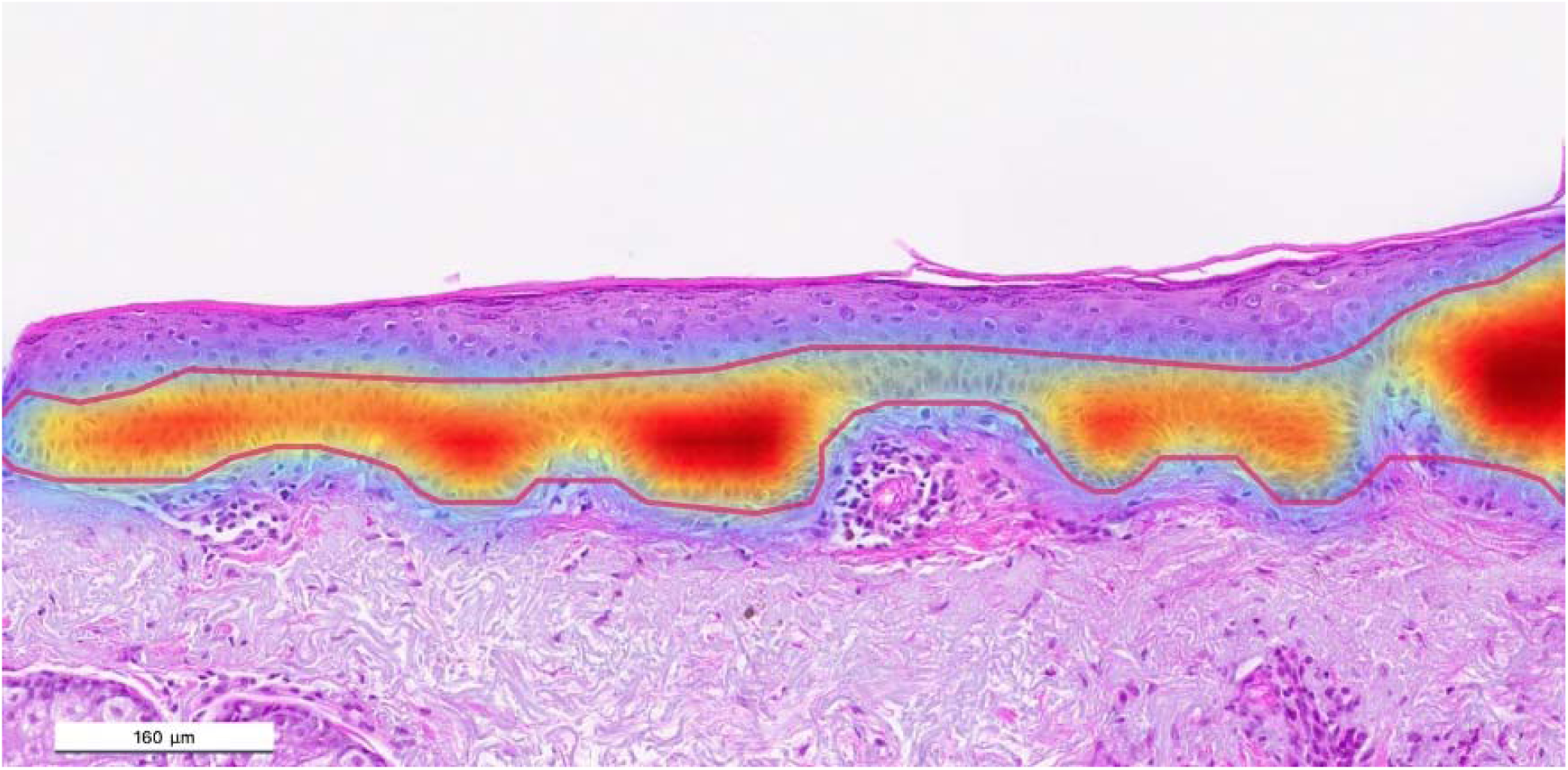
“Actinic Keratosis: WSI with heatmap”. WSI of an H&E slide with AK and the generated multi-color heatmap created by the DLM. Multi-color heat map s are proportional to the DLMs confidence from low to high confidence starting with light blue, then yellow, late r red, and finally dark red.

### 2.2 Training the DLM

The segmentation CNN was trained in a fully supervised way, i.e., the goal of the DLM was to match its output with the expert annotations. In total, 182 WSIs were used for the training of the DLM and 15% of the training data were randomly selected for validation. The pixel-wise expert annotations of AK regions were the target reference for the DLM. Additionally, a separate AI model resembling the architecture of the DLM used for AK was applied to create annotations of the epidermis. This separate model was developed in a previous project by authors of this study and trained for this purpose using hundreds of WSI with annotated epidermis. In this study, the epidermis model was employed to help the DLM quickly learn that AK originates from the epidermis, to allow it to focus on the differences between healthy and AK-affected epidermal regions. The DLM was trained for 60 epochs using the Adam optimizer [25] to minimize the focal loss [26]. Data augmentation, including random rotations, brightness, noise, elastic deformations, blur, contrast, and color variations were used to increase the robustness and performance of the DLM and to prevent the risk of overfitting. We ran the training on a GPU server with 4x Nvidia RTX A6000 (48GB) and implemented the DLM using the PyTorch framework [27]. The training progress was assessed and monitored on both the training and validation sets using the intersection over union (IoU) where higher values indicate a better match between the DLM and the dermatopathologists annotations.

### 2.3 Statistical analysis

For statistical analysis of the DLM performance, three different levels of granularity have been evaluated.

- WSI level: Assessment if AK was accurately detected by the DLM anywhere on the WSI dichotomously (with AK or without AK). Sensitivity, specificity, positive predictive value (PPV), negative predictive value (NPV), accuracy and balanced accuracy were calculated. Results on this level of granularity are congruent with the final diagnosis in dermatopathology.
- Patch level: Assessment if AK was accurately detected on each individual patch dichotomously (with AK or without AK). Sensitivity, specificity, PPV, NPV, accuracy and balanced accuracy were calculated. Results on this level of granularity evaluate which areas inside the WSI have been correctly or incorrectly classified.
- Pixel level: Assessment if AK was accurately detected on pixel level. The IoU is used as a performance metric without significance for the final diagnosis where sensitivity, specificity and accuracy have not been calculated. Instead, overconfidence and under-confidence were used to show the relative number of pixels that the DLM had annotated, with respect to the union area.

## 3 Results

### 3.1 Demographics

The dataset was congruent with the typical age and sex distribution of AK patients (Table 1) [28], [29]. 115 WSI were annotated by the dermatopathologists for evaluation of the trained

**Table 1.** “Database metrics”.

| Age |  |  | Gender |  |  |
| --- | --- | --- | --- | --- | --- |
|  | Train | Test |  | Train | Test |
| **Min** | 46,17 | 46,18 | **Male** | 108 | 57 |
| **Max** | 93,56 | 89,43 | **Female** | 58 | 48 |
| **Median** | 73,81 | 74,57 | **Male %** | 65,06% | 54,29% |
| **Average** | 71,18 | 73,1 | **Female %** | 34,94% | 45,71% |

AK DLM (105 with AK, 10 without AK). In total, 5815 non-overlapping image patches (each 1024×1024px) were extracted from the WSI and analyzed by the DLM. Here, 1770 patches had regions with AK that filled at least 1% of the total area of the patch (Table 2). In total, 29708 patches were extracted from the 182 WSIs. Here, 14004 had annotated regions with AK that filled at least 1% of the total area of the patch.

**Table 2.** “Composition of training and test data at WSI- and patch level”.

| **Dataset** | **Total Number of WSI** | **WSI with AK** | **WSI without AK** | **Total Number of patches** | **Patches with AK** | **Patches without AK** |
| --- | --- | --- | --- | --- | --- | --- |
| Train | 182 | 164 | 18 | 29708 | 14004 | 15704 |
| Test | 115 | 105 | 10 | 5815 | 1770 | 4045 |

### 3.2 WSI level

WSI with AK was detected by the DLM with a sensitivity of 100%. WSI without AK achieved a specificity of 90%. In WSI without AK, the DLM misclassified one area where an area of previously not noted flat SK was identified (Suppl. Figure 1). Overall, 99.13% accuracy was observed for the DLM on the WSI level (Table 3).

**Table 3.**
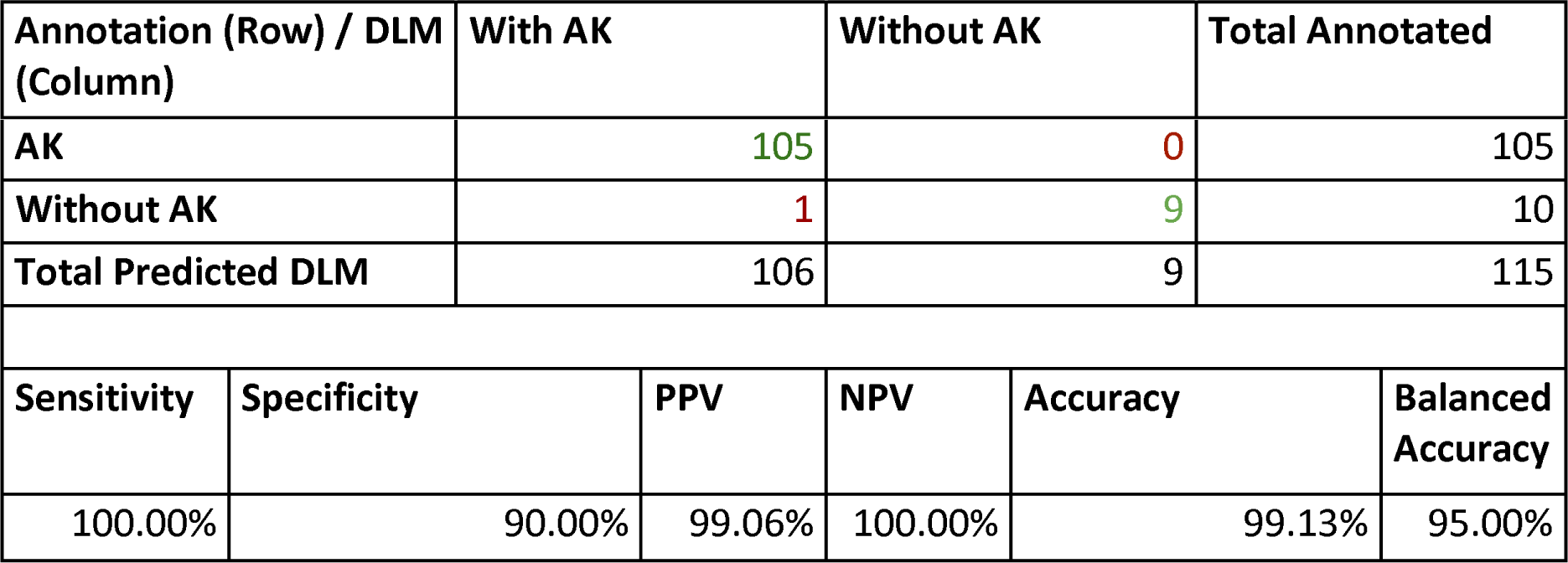
“Confusion Matrix and performance results on the test data: WSI level”.

### 3.3 Patch level

Patches with AK were detected by the DLM with a sensitivity of 98.87%. Patches without AK achieved a specificity of 99.09%. Overall, an accuracy of 99.02% was observed for the DLM on the patch level (Table 4). 20 patches were incorrectly classified as being without AK. They had been extracted from 16 different WSI (Supl. Figure 2). Conversely, 37 patches from 13 individual WSI were incorrectly classified as being with AK. Interestingly, one patch belonged to the WSI which was previously classified to contain only healthy epidermis and therefore was misclassified on the WSI level in Section 4.1. The others often included parts of hair follicles in 63.89% of the cases.

**Table 4.** “Confusion Matrix and performance results on the test data: Patch level”.

| Annotation (Row) / AI Model (Column) |  | AK | Without AK | Total Annotated |  |
| --- | --- | --- | --- | --- | --- |
| AK |  | 1749 | 20 | 1769 |  |
| Without AK |  | 37 | 4009 | 4046 |  |
| Total Predicted AI |  | 1786 | 4029 | 5815 |  |
| Sensitivity | Specificity | PPV | NPV | Accuracy | Balanced Accuracy |
| 98.87% | 99.09% | 97.93% | 99.50% | 99.02% | 98.98% |

### 3.4 Pixel level

The DLM also outputs a segmentation mask with a precise location of the AK region inside each image patch. This mask can be compared with the annotations created by the dermatopathologists. An IoU between the DLM and the dermatopathologists of 83.88% was observed. The DLM was overconfident for 12.18% of the union area, i.e., the DLM detected regions that were not annotated by the pathologists. Conversely, 3.94% of the union area was not detected by the DLM referred to as under confidence.

## 4 Discussion

The DLM achieved high sensitivity, specificity, and accuracy on all levels of granularity that were assessed. They were able to reliably detect AK in histopathological samples. The performance was evaluated on different granularity levels as results on the WSI level are more relevant for dermatopathology diagnostics, however patch and pixel levels are more interesting for technical assessment. DLMs have been shown to be accurate in the diagnosis of BCC, MB and SK [21] and this DLM has shown promising results for AK, a lesion with higher lesion intervariability (Figure 3). Explainable Artificial Intelligence (XAI) [30]–[32] is a novel approach which identifies parameters and their proportional influence on the training process. Several studies have shown a correlation between high intervariability in training data and learning performance for both machines and humans [33]–[36]. Histopathological diagnosis is based on distinct features which have been identified by humans due to recognition of repeating patterns [37]. DLMs also extract characteristic features and the number of features considered for final diagnosis could be higher for DLM compared with humans [38]. Additionally, the features extracted by DLM could be in a different dimension, as DLMs learn on the pixel level [39]. AK is a common in histopathology routine and incidence increases with age, posing challenges due to the aging population [29]. Near faultless performance on the WSI level verifies the realistic potential for using DLM in assisted dermatopathological diagnosis of AK. Key limitations of the study are that specimens were only collected from one histopathology lab and digitized with one scanner type. Variation in data from histopathology labs with different staining protocols or scanner types may influence the DLM performance, which was not analyzed in this study. Successful implementation of DLM software could improve the efficiency of daily clinical practice. DLMs could be used as screening tools to reduce workload. They also have potential to be developed to reduce the number of incorrect diagnoses due to human error. For example, in this study, the DLM detected epidermal alterations in one healthy tissue slide, inside an area of flat seborrheic keratosis. Research opportunities could also be facilitated with large volumes of automated analysis. The implementation of DLMs to daily dermatopathological practice faces many challenges. DLMs have been successfully implemented in histological analysis of the breast [40] and prostate by using applications to assist diagnosis [41], [42]. However, to date there is no commercially available software solution in dermatopathology. Three main barriers exist:

1. Economic burden for pathologists – the expense of slide scanners, data transfer infrastructure, digital workplace, servers, and data storage.
2. Merging DLM that analyze multiple frequent pathologies in usable software.
3. Capability of DLM to assist in pathology-specific diagnosis – DLMs are currently unable to provide information on variants or tumor metrics

**Figure 3.**
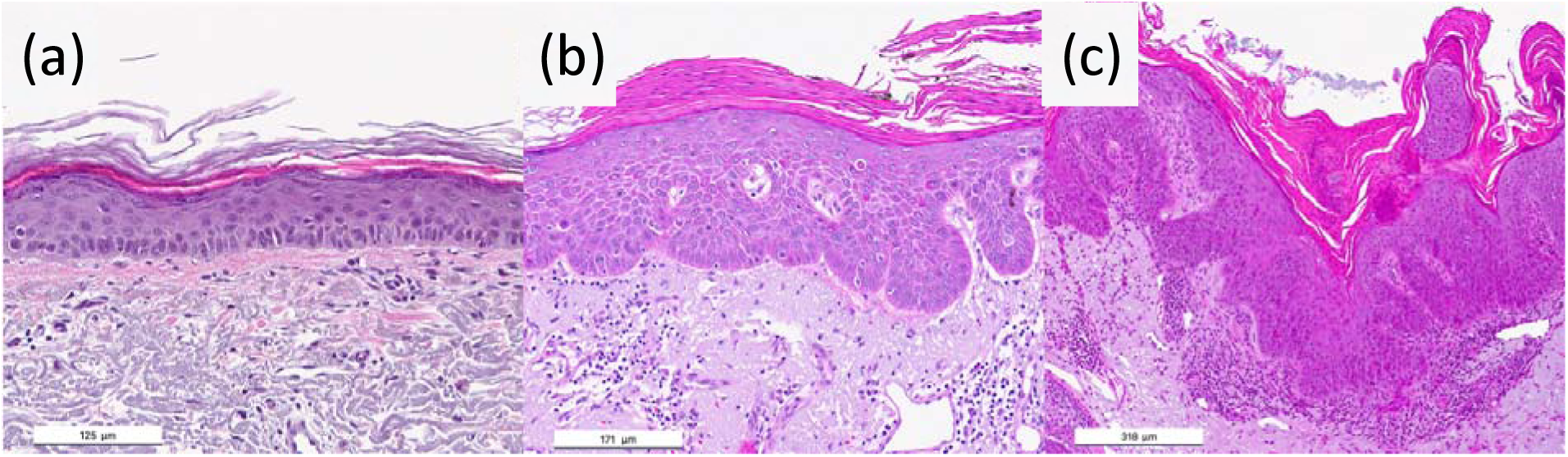
“Actinic keratosis: Histologic variability”. All slides are H&E. a) AK with atypical cells in the lower third of the epidermis (AK I) and no basal proliferation (PRO I) b) AK with ordered appearance and atypical cells throughout the whole epidermis width (AK III) as well as basal proliferation with the typical papillary sprouting (PRO III) c) AK with unordered appearance and atypical cells throughout the whole epidermis width (AK III) as well as extended basal proliferation also with papillary sprouting (PRO III).

This work uses AK as an example for research which must be done in the development of DLMs for assisted diagnostics in dermatopathology. Future work must be carried out to extend their capabilities. For AK, this will involve classifying upward and basal proliferation, and recognizing variants [24]. Furthermore, assessing the DLMs ability to differentiate between diagnoses and the behavior of DLMs in the presence of comorbidities must be evaluated. This DLM can reliably recognize AK in histopathological samples and supports the implementation of AI in real world dermatopathology practice. Given existing technical capabilities and advancements, DLMs could have a significant influence on dermatopathology routine in the future.

## Data Availability

Affiliates of the Centroderm Clinic1 have unlimited access to all clinical data used in this research work. Affiliates of the University of Bremen4 and aisencia5 have unlimited access to all technical data. Both affiliates share a pool of information for the collaboration. Clinical data of the included Actinic Keratoses are available in the published data set (Balkenhol, Julius; Schmidt, Maximillian; Schnauder, Tim; Langenhorst, Johannes; Le Clerc Arrastia, Jean; Otero Baguer, Daniel; Gilbert, Georgia; Schmitz, Lutz; Dirschka, Thomas (2023), Supplementary Data 1.0: The use of a deep learning model in the histopathological diagnosis of actinic keratosis: A case control accuracy study, Mendeley Data, V2, doi: 10.17632/2t5pg25vkh.2). Histopathological slides and annotations cannot be made publicly available. Access to view the slides and annotations can be given upon request to the authors.

https://data.mendeley.com/datasets/2t5pg25vkh/2

## Abbreviation List

AK: Actinic keratoses
ANN: Artificial neural networks
DNN: Deep neural networks
DLMs: Deep learning models
AI: Artificial intelligence
SK: Seborrheic keratosis
MB: Morbus Bowen
WSI: Whole slide image
PPV: Positive predictive value
NPV: Negative predictive value
XAI: Explainable Artificial Intelligence
IoU: Intersection over union

## 5 Acknowledgements

None

## 5 Legends of Supplementary Material

Supplementary Figure 1. “Actinic keratosis: DLM performance I”

a), b), c) Shows patch where AK was hallucinated in a hair follicle with red annotation by DLM. d), e), f) shows patch where the DLM missed AK with black annotation by the dermatopathologist. Moreover, b) and e) show single-color and c), f) show the multi-color heat map.

Supplementary Figure 2. “Actinic keratosis: DLM performance II”

a), b), c) Show misclassified patches in the healthy tissue cohort. d), e), f) Show correctly classified AK. a), d) show annotation by the DLM (red contour). b), d) show the single-color and e), f) the multi-color heat map where intensity is proportional to the DLMs confidence.

