## Supplementary Figure 1 + Supplementary Figure 2 for "The use of a deep learning model in the histopathological diagnosis of actinic keratosis: A case control accuracy study"

*Balkenhol, Julius; Schmidt, Maximillian; Schnauder, Tim; Langenhorst, Johannes; Le’Clerc Arrastia, Jean; Otero Baguer, Daniel; Gilbert, Georgia; Schmitz, Lutz; Dirschka, Thomas (2023), “Supplementary Data 1.0: The use of a deep learning model in the histopathological diagnosis of actinic keratosis: A case control accuracy study”, Mendeley Data, V1, doi: 10.17632/2t5pg25vkh.1*

***Mendeley Supplementary Figure 1 - Actinic keratosis: DLM performance I***

*a), b), c) Show misclassified patches in the healthy tissue cohort. d), e), f) show correctly classified AK. a), d) show annotation by the DLM (red contour). b), e) show the single-color and c), f) the multi-color heat map. Heat map intensity is proportional to the DLMs confidence.*

***Mendeley Supplementary Figure 2 - Actinic keratosis: DLM performance II***

*a), b), c) Show patch where AK was hallucinated in a hair follicle with red annotation by DLM. d), e), f) show patch where the DLM missed AK with black annotation by the dermatopathologist. b) and e) show single-color and c), f) show the multi-color heat map.*
